# Healthcare workers with mild / asymptomatic SARS-CoV-2 infection show T cell responses and neutralising antibodies after the first wave

**DOI:** 10.1101/2020.10.13.20211763

**Authors:** Catherine J. Reynolds, Leo Swadling, Joseph M. Gibbons, Corinna Pade, Melanie P. Jensen, Mariana O. Diniz, Nathalie M. Schmidt, David Butler, Oliver E. Amin, Sasha N. L. Bailey, Stephen Taylor, Jessica Jones, Meleri Jones, Wing-Yiu Jason Lee, Joshua Rosenheim, Aneesh Chandran, George Joy, Cecilia Di Genova, Nigel Temperton, Jonathan Lambourne, Teresa Cutino-Moguel, Mervyn Andiapen, Marianna Fontana, Angelique Smit, Amanda Semper, Ben O’Brien, Benjamin Chain, Tim Brooks, Charlotte Manisty, Thomas Treibel, James C Moon, COVIDsortium investigators, Mahdad Noursadeghi, COVIDsortium immune correlates network, Daniel M. Altmann, Mala K. Maini, Aine McKnight, Rosemary J. Boyton

## Abstract

Studies of adaptive immunity to SARS-CoV-2 include characterisation of lethal, severe and mild cases^1-8^. Understanding how long immunity lasts in people who have had mild or asymptomatic infection is crucial. Healthcare worker (HCW) cohorts exposed to and infected by SARS-CoV-2 during the early stages of the pandemic are an invaluable resource to study this question^9-14^. The UK COVIDsortium is a longitudinal, London hospital HCW cohort, followed from the time of UK lockdown^9,10^ ; weekly PCR, serology and symptom diaries allowed capture of asymptomatic infection around the time of onset, so duration of immunity could be tracked. Here, we conduct a cross-sectional, case-control, sub-study of 136 HCW at 16-18 weeks after UK lockdown, with 76 having had laboratory-confirmed SARS-CoV-2 mild or asymptomatic infection. Neutralising antibodies (nAb) were present in 90% of infected HCW sampled after the first wave; titres, likely to correlate with functional protection, were present in 66% at 16-18 weeks. T cell responses tended to be lower in asymptomatic infected HCW than those reporting case-definition symptoms of COVID-19, while nAb titres were maintained irrespective of symptoms. T cell and antibody responses were discordant. HCW lacking nAb also showed undetectable T cells to Spike protein but had T cells of other specificities. Our findings suggest that the majority of HCW with mild or asymptomatic SARS-CoV-2 infection carry nAb complemented by multi-specific T cell responses for at least 4 months after mild or asymptomatic SARS-CoV-2 infection.

---

The majority of people infected by SARS-CoV-2 have not been hospitalised and lack PCR confirmation of infection. A key concern is the extent to which immunity in mild or asymptomatic cases may confer protection from future infection^6,15-18^. The UK COVIDsortium recruited a cohort of 731 HCW at three London hospitals early in the pandemic at the time of UK lockdown (23^rd^ March 2020)^9,10^. HCW underwent longitudinal follow-up including weekly nasopharyngeal swabs for SARS-CoV-2 PCR, serum collection for antibody analysis and a self-reporting health questionnaire. 21.5% had laboratory confirmed infection and all were asymptomatic or had mild disease. We conducted a cross-sectional case-controlled sub-study (n=136) to analyse T cell and nAb immunity at 16-18 weeks after UK lockdown (Extended Data Table 1a). We collected samples from 76 HCW with laboratory-defined evidence of SARS-CoV-2 infection and 60 HCW matched for age, gender and ethnicity that were consistently SARS-CoV-2 PCR negative and serology negative. Here, we set out to investigate whether asymptomatic or mild infection with SARS-CoV-2 would confer specific nAb and T cell responses lasting beyond 4 months.

## SARS-CoV-2 multi-specific T cell response

A number of T cell studies investigating SARS-CoV-2 infection have described the presence of Th1 immunity^7^. We assessed SARS-CoV-2 T cell frequencies by IFNγ-ELISpot using three complementary approaches: whole protein^1^, mapped epitope peptide (MEP) pools^4^, and overlapping peptide (OLP) pools^3^ (Extended Data Table 2a, b). The use of whole protein allows assessment of CD4 T cell responses to naturally processed epitopes, whereas the MEP and OLP pools assessed a combination of CD4 and CD8 T cell responses directed against defined immunogenic regions and unbiased coverage of key viral proteins, respectively.

Analysing T cell responses to Spike and Nucleoprotein (NP) protein in HCW with mild or asymptomatic, laboratory confirmed infection, only 49% responded to Spike whereas significantly more (85%) responded to NP, showing a wide range of frequencies (Fig. 1a). Using MEP pools containing previously mapped immunogenic regions and offering good coverage for regional HLA genotypes^4^, responses of >80 SFC/10^6^ PBMC were found in 69% to peptide pools for Spike, NP, Membrane (M) and open reading frame (ORF)3a/7a, with the latter being at a significantly lower frequency. Eighty-seven percent of HCW had detectable T cell responses to these MEP pools (Fig. 1b). A third T cell stimulation platform used OLP pools spanning the whole of NP, M, and ORF3a, together with 15mers spanning immunogenic regions of Spike (Extended Data Fig. 1a); using this approach, we assessed multi-specificity and cumulative SARS-CoV-2-specific T cell frequencies. This indicated a wide range of cumulative T cell response frequencies, from zero to >1000 SFC/10^6^ PBMC, with 86% showing a detectable T cell response (Fig. 1c). Of note with both the MEP and OLP platforms, responses to ORF3a/7a or ORF3a respectively were significantly lower than other antigens (Fig. 1b; Extended Data Fig. 1a). Although T cell responses to individual regions were relatively weak, their cumulative frequencies across all pools tested were similar in magnitude to that of T cells directed against a pool of well-described CD8 epitopes from influenza, EBV and CMV assessed in parallel in the same donors (Extended Data Fig. 1a), and comparable to frequencies found against SARS-CoV-1 pools following SARS infection^19^

**Figure 1.**
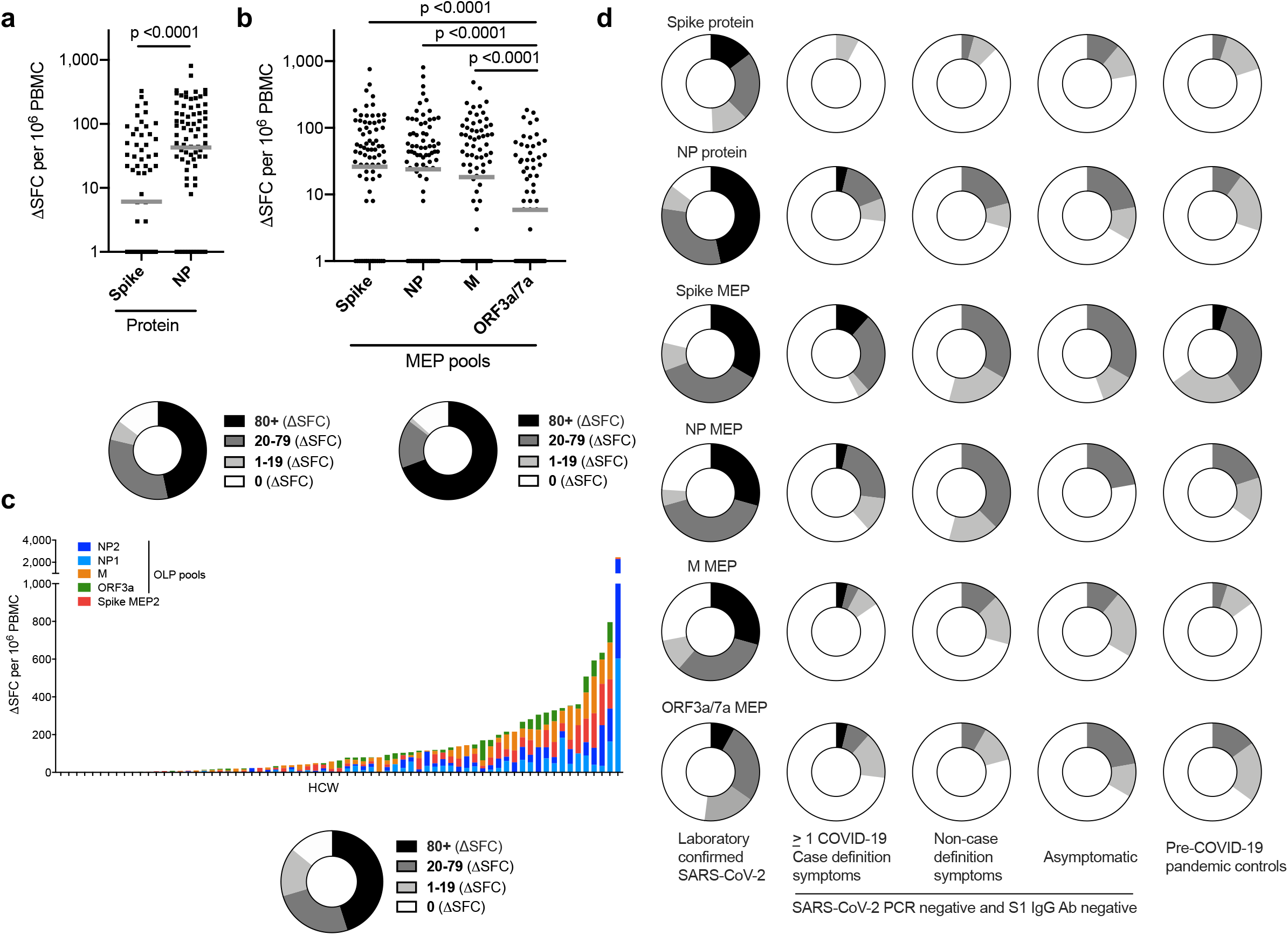
T cell responses to SARS-CoV-2 antigens in HCW (laboratory-confirmed COVID-19) at 16-18 weeks after UK lockdown: **a-c)** Magnitude of T cell response and proportion of HCW with a summed T cell response within the given ranges (0, 1-19, 20-79, ≥80 ΔSFC/106 PBMC). **a**, Spike and NP protein (n =75), **b**) mapped epitope peptide (MEP; n = 75) and **c**) overlapping peptide (OLP) pools (n =71, ordered by cumulative magnitude) in HCW with laboratory-confirmed SARS-CoV-2 infection (n = 75). **d)** Proportion of HCW with a T cell response to SARS-CoV-2 individual proteins or peptide pools within given ranges (0, 1-19, 20-79, ≥80 ΔSFC/106 PBMC) in the following groups: HCW cohort with laboratory-confirmed infection (n = 75); HCW cohort with no laboratory-confirmed infection but with one or more case-definition symptoms (n = 26), non-case-definition symptoms (n = 24) or asymptomatic (n = 9); pre-COVID-19 pandemic control cohort 1 (n = 20). **a-b)** Bars at geomean. **a)** Wilcoxon matched-pairs signed rank test. **b)** Friedman multiple comparisons ANOVA with Dunn’s correction. Ab, antibody; HCW, health care workers; M, Membrane; ORF, open reading frame; NP, Nucleoprotein; S1, Spike subunit 1; SFC, spot forming cells per 10^6^ PBMC.

Responses to Spike, NP protein and Spike, NP and M MEP were significantly higher frequency in HCW with laboratory confirmed SARS-CoV-2 infection than those in the matched group without laboratory evidence of infection (Fig.1d, Extended Data Fig. 1, c-d). For example, 85% and 49% of HCW with laboratory confirmed SARS-CoV-2 had T cell responses to NP and Spike protein respectively, compared with 29% and 12% of SARS-CoV-2 PCR negative and S1 IgG negative HCW (p<0.0001) (Fig.1d). T cell recognition of these stimuli in HCW without evidence of infection, irrespective of reported COVID-19-like symptoms, was similar to that seen in pre-COVID-19 pandemic controls (Fig. 1d, Extended Data Fig. 1c, d; Extended Data Table 1a, b). The OLP pools (utilising increased cell numbers) showed detectable T cell responses in the PCR negative, S1 IgG negative HCW group (Extended Data Fig. 1b). With every T cell stimulation approach tested, responses were also seen in a proportion of pre-pandemic controls. Epitope mapping studies will be required to investigate possible cross-reactive components of these responses with other human coronaviruses as other studies have highlighted^2,3,20^ and to assess the impact of any such cross-reactivity on disease outcome, whether positive or negative^21,22^.

In addition to IFNγ SFC, we explored other cytokines indicative of non-Th1 subset polarisation by screening supernatants from Spike and NP protein-stimulated ELISpots; they showed no evidence of IL-4, 5, 13, 17 or 23 (Extended Data Fig. 1e).

In line with previous observations of SARS-CoV-2 T cells and ageing^23^, T cell responses in HCW (n=75) with laboratory confirmed SARS-CoV-2 correlated with age. There was a correlation with increasing age and T cell responses against Spike MEP2, NP1 OLP and ORF3a/7a MEP (Extended Data Fig. 2a-d). Broken down by age and gender, T cell immunity to Spike increased with age in males (Spike protein; r=0.522, p=0.006) (Extended Data Fig. 2e). We found no differences in T cell responses associated with ethnicity (Extended Data Fig. 3a,b). T cell immunity to M MEP, ORF3a/7a MEP and ORF3a OLP was higher in males compared to females (Extended data Fig. 3c,d).

### Neutralising antibodies to SARS-CoV-2 at 16-18 weeks

The majority of HCW in this cohort with laboratory confirmed SARS-CoV-2 infection had detectable S1 IgG and/or NP IgG/IgM (97%) during follow-up. Peak antibody level during 16-18 week follow-up (Fig2a) was considered to be a useful marker of humoral immune activation in each HCW. Some studies of nAb responses in severe, mild and asymptomatic disease have highlighted rapid waning of nAb within weeks^14-16, 24^, with others finding a more sustained neutralising response^25-27^. We analysed the nAb response in HCW at 16-18 weeks after UK lockdown and found that 90% could neutralise pseudotype virus. There was a range of nAb titres detectable, with 66% having an IC50 titre of >200 (Fig. 2b-c), a correlate of protection in SARS-CoV-2 based on viral challenge in macaque studies^28^. Ten percent of HCW with laboratory-confirmed SARS-CoV-2 infection demonstrated no detectable neutralising response (Fig. 2b-c). Typical nAb profiles in the high (IC50 ≥200), low (IC50 50-199) and none (IC50 ≤49) categories are shown (Extended Data, Fig 4a). The nAb response positively correlated with peak S1 IgG and peak NP IgG/IgM (Fig. 2d). Peak S1 IgG tended to be lower in those reporting non-case defining symptoms and those who were asymptomatic compared to those with case-definition symptoms (Fig. 2e). However, the nAb IC50 titre at 16-18 weeks after lockdown was maintained at a similar level across these three symptom groups (Fig. 2f). Eighty-eight percent of HCW aged ≥50y developed nAb at an IC50 of >200 compared with 59% of younger HCW aged 24-49y; p=0.041 (Fig. 2g). Peak S1 IgG Ab and nAb IC50 increased with age in females (Fig 2h). We looked in more detail at comparative features of infected individuals in the HCW cohort who did or did not show a nAb response at 16-18 weeks (Extended Data, Fig. 4b-d); we cannot discount the possibility that these individuals may have shown an earlier response that had waned by 16-18 weeks. These 7 HCW with no nAbs spanned an age-range of 26-53y and tended to be at the lower end of the HCW age-range. Although this sub-study was not powered to investigate stratified demographic differences, we looked at features such as gender, ethnicity, clinical role or location, use of personal protective equipment (PPE) or symptom profile and found no difference between those that made nAb and those that did not, though there was a trend to more male non-neutralisers.

**Figure 2.**
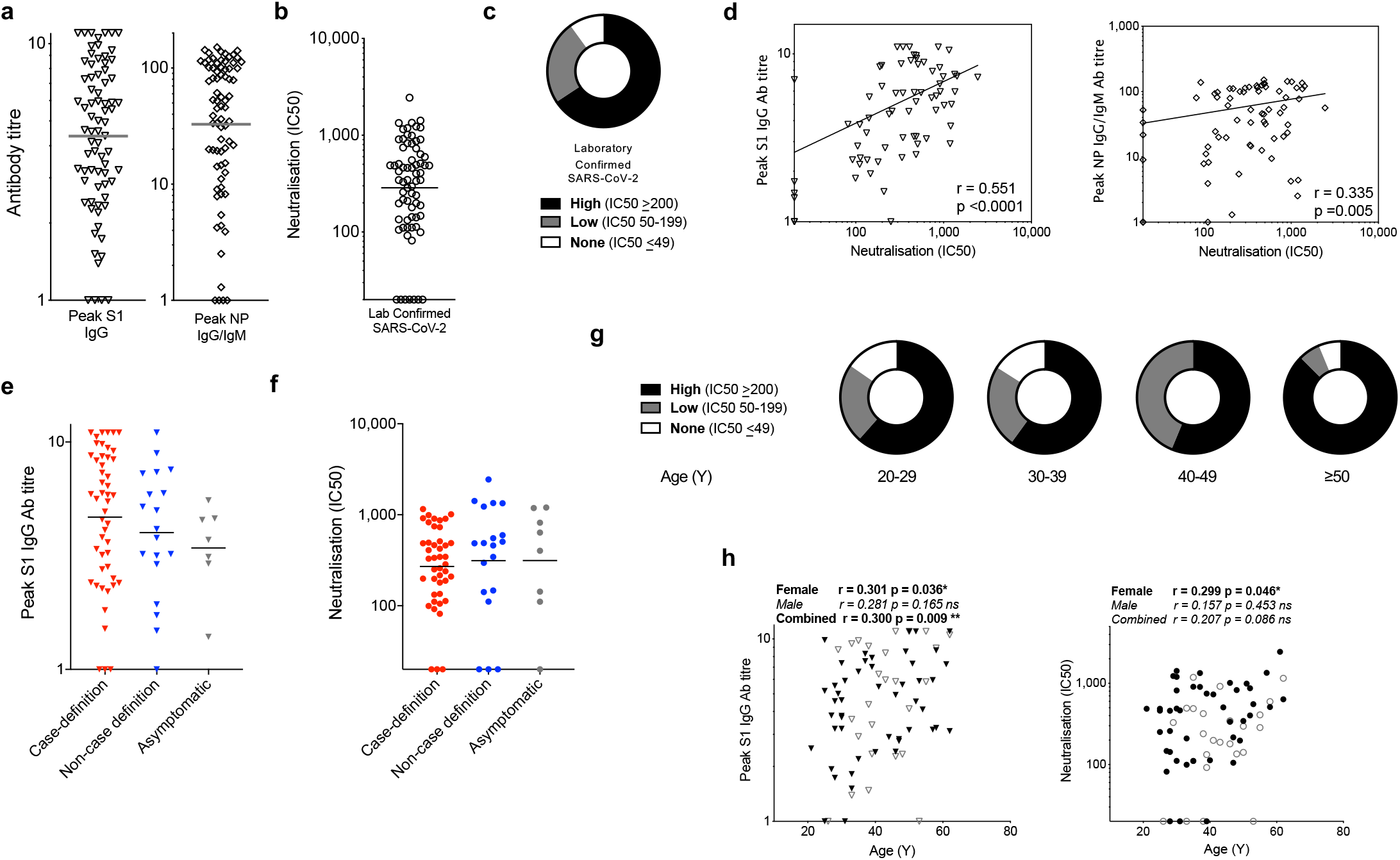
nAb responses to SARS-CoV-2 antigens in HCW (laboratory-confirmed COVID-19) at 16-18 weeks after UK lockdown: **a)** Peak S1 IgG antibody titre and peak NP IgG/IgM Ab titre across the study period in HCW with laboratory-confirmed SARS-CoV-2 infection (n = 70). **b)** The distribution of nAb (IC50) titres across the cohort of HCW with laboratory-confirmed infection and **c)** The proportion of HCW with an undetectable (0-49), low (50-199) or high (200+) nAb titre (IC50). **d)** Correlation between peak S1 IgG Ab titre (left) or the peak NP IgG/IgM Ab titre (right) and nAb titre (IC50) in HCW with laboratory-confirmed SARS-CoV-2 infection. **e-f**) Peak S1 IgG Ab titre **e**) and nAb titre (IC50) **f**) in HCW with laboratory-confirmed infection, stratified by symptom group: ≥1 COVID-19 case-definition symptoms (Red), non-case definition symptoms (Blue) or asymptomatic (Grey) throughout trial and within 3-months of trial initiation. **g)** The proportion of HCW with an undetectable (0-49), low (50-199) or high (200+) nAb titre (IC50) within specified age ranges; 20-29 Y (n = 13), 30-39 Y (n = 25), 40-49 Y (n = 16) and ≥50 Y (n = 16). **h)** Correlations of age vs. peak S1 IgG Ab titre (left) and neutralising antibody titre (IC50; right) in HCW with laboratory-confirmed SARS-CoV-2 infection separated by gender (female, black symbols; male, open symbols). **d, h)** Spearman’s rank correlation, least squares log-log lines shown. **a-b, e-f)** bars at geomean. **e, f)** Kruskall Wallis multiple comparison ANOVA with Dunn’s correction, not significant. Ab, antibody; nAb, neutralising antibody; S1, Spike subunit 1; SFC, spot forming cells per 10^6^ PBMC; Y, years.

### T cell and nAb responses are sometimes discordant

To better understand complementarity between nAb and T cells, we next compared the T cell, S1 IgG and nAb responses in individual HCW. T cell responses to Spike and NP protein correlated with peak S1 IgG titre, but with weak correlation coefficients partly attributable to lack of T cell responses in some HCW with positive antibody titres to Spike and NP (Fig. 3a; blue box in Extended Data Figs. 5a, d). Correlations between peak NP IgG/IgM titre and T cell responses to Spike and NP protein showed similar results (Extended Data Fig. 6a). Just over half HCW were discordant for T cell and S1 IgG responses, making no T cell response to Spike protein and 15% made no T cell response to NP(Extended Data Fig. 5a-f). While we found no differences in terms of age, gender, ethnicity, symptom profile, clinical role or PPE use, there tended to be more non-responders among Black, Asian and minority ethnic (BAME) HCW.

**Figure 3.**
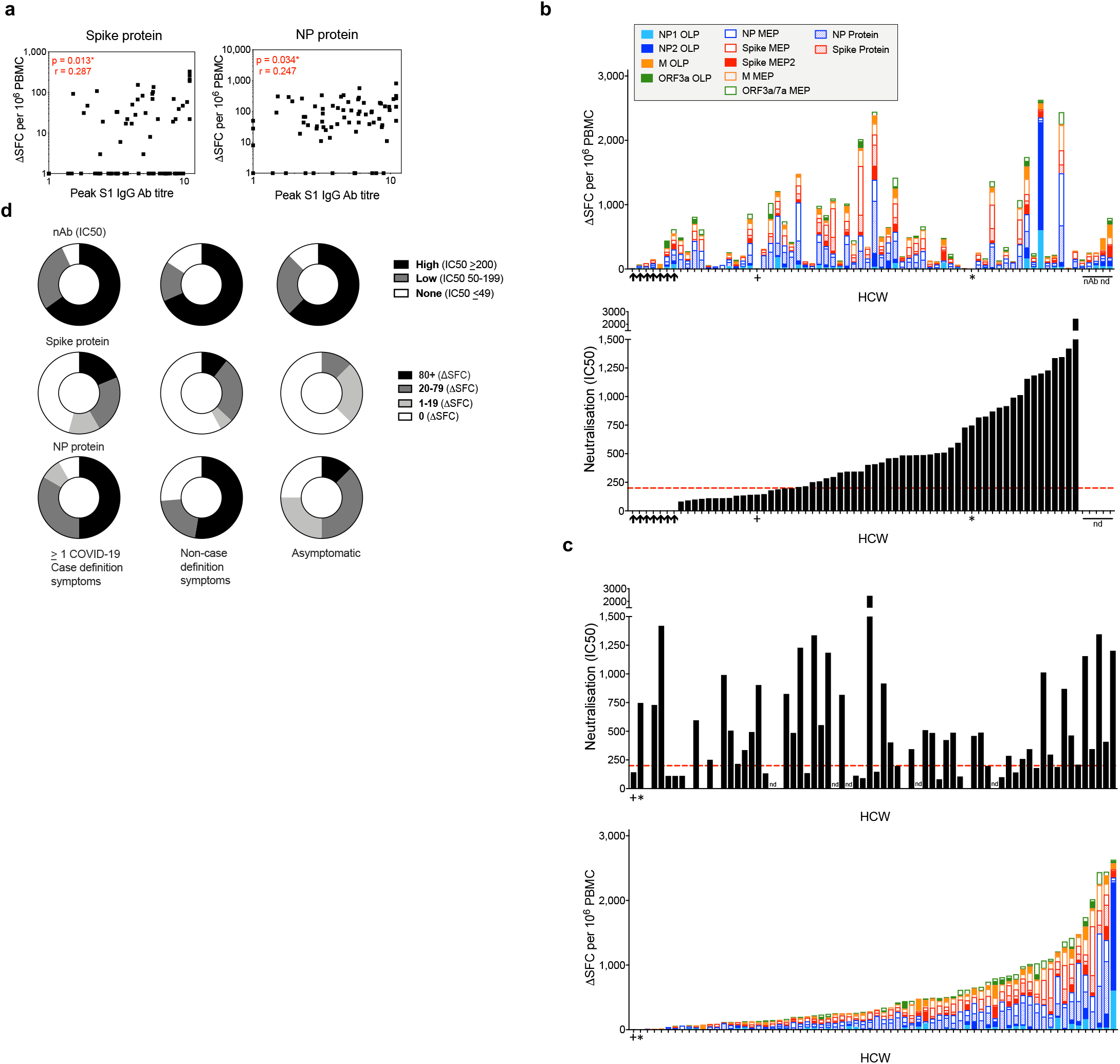
Concordant and discordant T cell and nAb responses in HCW (laboratory-confirmed COVID-19) at 16-18 weeks after UK lockdown. **a)** Correlations between the peak S1 IgG Ab titre and T cell responses to Spike protein (left) or NP protein (right) in HCW with laboratory-confirmed SARS-CoV-2 infection (n = 75). **b)** Top panel; Cumulative magnitude of the T cell response to Spike and NP proteins, mapped epitope peptide (MEP and MEP2) panels and overlapping peptide (OLP) panels (top panel) ordered by increasing magnitude of nAb response (bottom panel) or **c)** Magnitude of nAb response (top panel) ordered by increasing cumulative magnitude of T cell response to Spike and NP proteins, MEP/MEP2 panels and OLP panels (bottom panel) in HCW with laboratory-confirmed SARS-CoV-2 infection (n = 70). HCW with no nAb (IC50 titre less than 50) are indicated by black arrows. + and * denote individuals with no T cell response to any protein or peptide pool. **d)** Proportion of HCW with a nAb titre (IC50) or T cell response to Spike and NP proteins within given ranges stratified by symptom group; ≥1 COVID-19 case definition symptoms (n = 43 or 48), non-case definition symptoms (n = 19) or asymptomatic (n = 8) **a)** Spearman’s rank correlation. HCW, health care workers; M, Membrane; ORF, open reading frame; nAb, neutralising antibody; NP, Nucleoprotein; S1, Spike subunit 1; SFC, spot forming cells per 106 PBMC.

In Fig. 2b, we showed that 10% of infected HCW lacked detectable nAb at 16-18 weeks after UK lockdown. To understand the complementarity between T cell and nAb responses in individual HCW, we analysed responses of all HCW ranked either by nAb IC50 titre or cumulative T cell response. We first arrayed HCW responses ranked by magnitude of nAb response (Fig. 3b). Neutralisation IC50 values for all HCW were plotted in relation to our indicative, protective cut-off value of >200 (dotted horizontal red line in lower panel). HCW lacking detectable nAb are indicated by 7 black arrows on the left. Their cumulative T cell response frequencies against viral antigens are shown in the panel above and are relatively low. Relating lack of nAbs to T cell responses to different specific antigens we show that none of the 7 HCW without detectable nAb make a T cell response to Spike protein (Extended data Fig 6b). The addition of data from Spike MEP pools (potentially encompassing CD8 responses as well) revealed low T cell responses to Spike in 4/7 HCW lacking a nAb response (Extended data Fig 6c). Exploring T cell responses to NP protein and NP, M, and ORF3a/7a MEP pools showed 5/7 HCW without detectable nAb making a T cell response (Extended data Fig 6d). Furthermore, there were OLP T cell responses in 5/7 HCW lacking nAb (Extended data Fig 6e). Thus HCW lacking nAb tend to lack responses to Spike while maintaining low frequency T cells to other specificites.

Examining the converse, we then arrayed HCW responses ranked by magnitude of cumulative T cell response (Fig 3c). From this plot, HCW with the lowest cumulative T cell response (to the left of the plot) have a range of nAb responses from none to >200 IC50. One young, asymptomatic, female HCW with a good peak S1 IgG titre had no T cell response to any antigens tested but made nAbs with a titre of 143, which may be insufficient for functional protection (Fig 3b,c indicated by +). Another female HCW with a good S1 IgG titre, also had no T cell response to any antigens tested, but made nAbs with a titre of 747 (Fig 3b,c indicated by *). Assessing T cell responses ranked simply on the basis of presence or absence of recognition of proteins and pools (rather than magnitude of response) indicates that those lacking a nAb response (black arrows) showed T cell responses against 2 to 5 antigens (Extended data Fig 6f). Taken together, the data show discordance of nAb and T cell responses in individual HCW.

Of the 76 HCW studied with mild or asymptomatic laboratory-confirmed SARS-CoV-2 infection, 64% had one or more case-defining symptoms, 25% had non-case-defining symptoms and 11% were asymptomatic. Looking at T cell immunity and nAb levels across these symptom-stratified groups at 16-18 weeks, T cell responses tended to be higher in infected HCW with case defined symptoms. Responses to M MEP and ORF3a OLP were significantly higher in HCW reporting case definition symptoms than those that were asymptomatic (Extended Data Fig. 7a, b).

Importantly, there was no significant fall in nAb titres across case-defining, non-case-defining symptoms and asymptomatic HCW groups (Fig. 2d) with 65%, 68% and 63% respectively showing an IC50 >200. Thus, the majority of HCW with laboratory-confirmed infection have a detectable (and likely protective) level of nAbs at 16-18 weeks, whilst the 10% who lack detectable nAb have fewer T cells directed against Spike but can show reactivity to other regions of the viral proteome.

## Discussion

Much debate has focused on the possibility that the Ab response to SARS-CoV-2 may be short-lived, while T cell recognition may be strong, durable, and more common ^14,16,17,23,27,29^. Mild or asymptomatic infection are very common but are not usually diagnosed contemporaneously, making assessment of the durability of immunity in this common group challenging. Here we describe a cross-sectional study of an exposed HCW cohort at 16-18 weeks after UK lockdown who had mild or asymptomatic infection picked up by repeated PCR and serological testing. This cohort shows variable T cell responses across the viral proteome sampled, with only two HCW with lab-confirmed COVID-19 showing no detectable T cell response across all the platforms tested. In this study, 90% of HCW with asymptomatic or mild COVID-19 had nAb at 16-18 weeks after UK lockdown and 66% had titres >200. In light of some reports of rapid waning of nAbs this result was surprising^15-17,24,29^. Here we show a complex pattern of T cell and nAb responses for individual HCW. Analysis of nAbs shows that the majority of laboratory confirmed SARS-CoV-2 infected HCW with no symptoms or only mild disease had relatively high nAb IC50 at 16-18 weeks after UK lockdown. These IC50s were in the same range as those defined as conferring functional protection in macaque challenge studies^28^. In terms of neutralisation observations in humans, a study of SARS-CoV-2 susceptibility during an outbreak on a fishing vessel indicated a lack of infection in those showing a prior nAb titre (IC50) >1/160^30^. In infection by SARS-CoV-1, nAbs are often lost by 1-2 years after infection^19,31^, whereas T cell responses can persist for up to 17y after SARS-CoV-1^3^. Longitudinal follow-up of nAb versus T cell kinetics in the COVIDsortium cohort will illuminate T cell and nAb trajectories over time.

In terms of severe COVID-19 risk, two of the strongest factors identified have been gender and age^32^. We found a positive correlation between both S1 IgG Ab level and nAb IC50 with age in female study participants. Other observations have suggested a higher T cell response to mitogens in females with acute hospitalised COVID-19^33^ ; we observed higher memory T cell responses to Spike antigen in older males at 16-18 weeks after UK lockdown. Thus, in this asymptomatic/mild cohort of HCW, the nAb response increases significantly with age in females, while it is the T cell response that increases significantly with age in males.

A cautionary note about the ephemeral nature of adaptive immunity to coronaviruses comes from data for annual reinfections with the four seasonal coronaviruses and emerging data for reinfection by SARS-CoV-2^34,35^. Some studies have raised concern about the durability of serum antibodies and B cell memory, with data pointing towards impaired germinal centre reactions in severe acute COVID-19^29^. Other studies have focused on the potential for rapid waning of nAb after mild SARS-CoV-2 infection^14,15^. However we find nAb detectable in the majority of HCW sampled 16-18 weeks after mild/asymptomatic infection. Some T cell data indicates that even asymptomatic people and household contacts develop low-frequency T cell responses, in line with results from the HCW without laboratory confirmed infection using one of our platforms with higher T cell numbers^6^. We show here that infected HCW can display highly heterogeneous T cell recognition of epitopes from diverse SARS-CoV-2 structural and non-structural proteins, but it is not yet possible to decode the differential impacts of these responses for protection. Analysis of T cell response repertoire in convalescent, hospitalised COVID-19 patients argues that breadth of T cell response is a marker of mild disease^36^.

In summary, this study of HCW with laboratory confirmed SARS-CoV-2 infection finds that in the majority of these working adults there is immunity at 16-18 weeks comprising nAb (often at a level likely to protect), usually complemented by multi-specific T cell responses. Understanding protective immunity in the population will require simultaneous scrutiny of T cell and antibody responses.

## Data Availability

All data has been reported in the manuscript, Figures and Extended Data Figures and Tables.

## Data Availability

All data has been reported in the manuscript, Figures and Extended Data Figures and Tables.

## Methods

### Ethics statement

The COVIDsortium Healthcare Workers bioresource was approved by the ethical committee of UK National Research Ethics Service (20/SC/0149) and registered on ClinicalTrials.gov (NCT04318314). The study conformed to the principles of the Helsinki Declaration, and all subjects gave written informed consent.

Pre-pandemic healthy donor samples were collected and cryopreserved before October 2019. Pre-pandemic cohort 1 and 2 samples were recruited under ethics numbers 17/LO/0800 and 11/LO/0421 respectively.

### COVIDsortium Healthcare Worker Participants

Adult HCW (>18 years old) from a range of clinical settings who self-declared as fit to attend work were invited to participate via local advertisement of the project (see https://covid-consortium.com). Full study details of the bioresource (participant screening, study design, sample collection, and sample processing) have been previously published^10^.

A cohort of 400 HCW was initially recruited from St Bartholomew’s Hospital, London, in the week of UK lockdown (23^rd^-31^st^ March 2020). All participants were asymptomatic and self-declared fit to attend work in hospital. Recruitment was extended (27^th^ April-7^th^ May 2020) to include 331 additional participants from multiple sites: St Bartholomew’s Hospital (n=101 additional), NHS Nightingale Hospital (n=10), and Royal Free NHS Hospital Trust (n=220).

A prospective, observational, longitudinal cohort design was used and consisted of questionnaires exploring demographic, clinical and exposure risks, and sample collection at baseline and weekly follow-up for 15w from the start of each cohort. Participants were asked to provide details and timing of symptoms in the 3 months prior to baseline, and for those who were unable to attend follow-up visits (due to shift rostering, annual leave or self-isolation), the reason for non-attendance was collected, to ensure capture of information regarding isolation due to participant symptoms or household contacts. On return from self-isolation with symptoms, convalescent samples were collected. Further follow-ups at 6 and 12 months are planned.

Complete details of the sampling protocol have been previously published^10^. Initial analysis of samples for determining infection with SARS-CoV-2 included: nasal RNA stabilizing swabs baseline and weekly with reverse transcriptase polymerase chain reaction (RT-PCR): Roche cobas® SARS-CoV-2 test; Ab testing baseline and weekly: IgG Ab assay to spike protein S1 antigen, (EUROIMMUN Anti-SARS-CoV-2 enzyme-linked immunosorbent assay [ELISA]); and anti-nucleocapsid total antibody assay (ROCHE Elecsys Anti-SARS-CoV-2 electrochemiluminescence immunoassay [ECLIA]). Antibody ratios ≥ 1.1 were considered test positive for the EUROIMMUN SARS-CoV-2 ELISA and >1 was considered test positive for the ROCHE Elecsys anti-SARS-CoV-2 ECLIA following published Public Health England (PHE) evaluations.

Evaluation of Roche Elecsys Anti-SARS-CoV-2 serology assay for the detection of anti-SARS-CoV-2 antibodies. PHE, UK 110620. [https://assets.publishing.service.gov.uk/government/uploads/system/uploads/attachment_data/file/891598/Evaluation_of_Roche_Elecsys_anti_SARS_CoV_2_PHE_200610_v8.1_FINAL.pdf]

Evaluation of Euroimmun Anti-SARS-CoV-2 ELISA (IgG) serology assay for the detection of antibodies. PHE, UK [https://assets.publishing.service.gov.uk/government/uploads/system/uploads/attachment_data/file/893433/Evaluation_of_Euroimmun_SARS_CoV_2_ELISA_IgG1_.pdf]

At baseline, information relating to demographics and exposures was collected via a standardised questionnaire. Mean age of the cohort (n=731) was 38±11 years; 33% are male, 31% nurses, 20% doctors, and 19% work in intensive care units. COVID-19-associated risk factors were: 37% Black, Asian or minority ethnicities (BAME); 18% smokers; 13% obesity; 11% asthma; 7% hypertension and 2% diabetes mellitus^10^. At weekly follow-up visits information relating to symptom burden was recorded using a standardised questionnaire. Symptoms were classified as follows: ‘case-defining’ (fever, new continuous dry cough or a new loss of taste or smell), ‘non-case-defining’ (specific symptoms other than case-defining symptoms, or unspecified symptoms), or asymptomatic (no symptoms reported).

Case definition for coronavirus disease 2019 (COVID-19), as of 29 May 2020 European Centre for Disease Prevention and Control [https://www.ecdc.europa.eu/en/covid-19/surveillance/case-definition]

A total of 731 HCW underwent 16 weeks of serial assessment (attending unless ill, self-isolating, on holiday, or redeployed). Across the main study cohort, 48 participants had positive RT-PCR results with 157 (21.5%) seropositive participants. Infections were asymptomatic or mild with only two hospital admissions (neither requiring intensive care admission, both discharged well). The cohort therefore represents working age community COVID-19 rather than hospitalised COVID-19.

The cross-sectional case controlled sub-study (n=136) collected samples at 16-18 weeks after UK lockdown (Extended Data Table 1a, Extended Data Fig. 8). The cross-sectional case controlled sub-study included 76 HCW (mean age 41y, 36% male) with laboratory defined evidence of SARS-CoV-2 either by SARS-CoV-2 positive PCR and/or positive for spike IgG (Euroimmun ELISA)/ NP IgG/IgM antibody (Roche Elecsys). Fifty-seven percent reported one or more case defining COVID-19 symptoms. Twenty-four percent reported non-case defining symptoms and 19% were asymptomatic at baseline, during 16-week follow-up or in the 3 months prior to baseline. A second age, gender, and ethnicity matched subgroup of sixty HCW were recruited (mean age 39y, 37% male) who were SARS-CoV-2 PCR negative and negative for Spike IgG (Euroimmun ELISA) and NP IgG/IgM antibody (Roche Elecsys) tests throughout the 16-week follow-up. However, forty-four percent reported one or more case defining COVID-19 symptoms, 41% non-case defining symptoms and 15% were asymptomatic at baseline, during 16-week follow-up and in the 3 months prior to baseline.

### Isolation of PBMC

Peripheral blood mononuclear cells (PBMC) were isolated from heparinized blood samples using Pancoll (Pan Biotech) or Histopaque®-1077 Hybri-Max™ (Sigma-Aldrich) density gradient centrifugation in SepMate™ tubes (Stemcell) according to the manufacturer specifications. Isolated PBMCs were cryopreserved in fetal calf serum containing 10% DMSO and stored in liquid nitrogen.

### Isolation of serum

Whole blood samples were collected in SST vacutainers (VACUETTE® #455092) with inert polymer gel for serum separation and clot activator coating. After centrifugation at 1000 X g for 10 minutes at room temperature, serum layer was aliquoted and stored at -80^□^C for specific SARS-CoV-2 Ab titre detection by ELISA and for SARS-CoV-2 Spike pseudotyped virus neutralisation assays.

### SARS-CoV-2 specific Ab titre

Anti-SARS-CoV-2 S1 IgG ELISA (EUROIMMUN) was performed on a Stratec Biomedical Gemini automated ELISA platform as described. The optical density was detected at 450nm, and a ratio of the reading of each sample to the reading of the calibrator included in the kit was calculated for each sample. An OD ratio of ≥1.1 was interpreted as positive for S1 antibodies by manufacturer’s recommendation.

Elecsys® Anti-SARS-CoV-2 nucleocapsid total Ab ELISA (ROCHE) was performed as described. Results are reported as numeric values in the form of a cut-off index (COI; signal sample/cut-off) as well as in the form of qualitative results non-reactive (COI < 1.0; negative) and reactive (COI ≥ 1.0; positive).

### Recombinant proteins

The SARS-CoV-2 S1 Spike antigen and Nucleoprotein was obtained from the Centre for AIDS Reagents (CFAR), National Institute for Biological Standards and Control (NIBSC), UK: SARS-CoV-2 Nucleoprotein and S1 Spike antigen from Dr Peter Cherepanov, Francis Crick Institute, UK.

### Mapped epitope pools (MEP)

Pools of 13-20mer peptides based on the protein sequences of SARS-CoV-2 S1 (Spike), nucleoprotein (NP), membrane (M) and open reading frames 3a and 7a (ORF3a/7a) described previously were synthesized^4^ (GL Biochem Shanghai Ltd, China). To stimulate PBMC, separate pools of sequences for Spike (18 peptides), NP (10 peptides), M (6 peptides) and ORF3a/7a (7 peptides) were used, see Extended Data Table 2a). A second mapped epitope pool of SARS-CoV-2 S1 peptides (Spike MEP2) based on alignment of all sequences of published SARS-CoV-1 epitopes (www.iedb.org; search criteria: positive assays only, T cells assays, host: human) with the Spike-SARS-CoV-2 sequence and 15-mer peptides synthesised to cover the homologous sequences. In addition, we synthesised 15-mer peptides covering the predicted SARS-CoV-2 Spike epitopes^3^ to give a total of 55 peptides in this pool (Spike MEP2), see Extended Data Table 2b

### Overlapping peptide pools (OLP)

15-mer peptides overlapping by 10 amino acids spanning the entire protein sequence of SARS-CoV-2 nucleoprotein (NP), Membrane (M) and ORF3a were synthesized (GL Biochem Shanghai Ltd; see Extended Data Tables 2b). To stimulate PBMC, the peptides were divided into 4 pools covering NP (NP-1, NP-2, 41 peptides each), M (43 peptides), and ORF3a (53 peptides).

### IFNγ-ELISpot Assay

Unless otherwise stated, culture medium for human T cells was sterile 0.22µM filtered RPMI medium (GibcoBRL) supplemented with 10% by volume heat inactivated (1h, 64°C) fetal calf serum (FCS; Hyclone, and 1% by volume 100x penicillin and streptomycin solution (GibcoBRL).

For experiments involving T cell stimulation with proteins or MEP peptide pools, pre-coated ELISpot plates (Mabtech 3420-2APT) were washed x4 with sterile PBS and were blocked with R10 for 1h at room temperature. 200,000 PBMC were seeded in R10/well and were stimulated for 18-22h at 37°C with 5%CO_2_ with SARS-CoV-2 recombinant proteins (10µg/ml) or MEP pools (10µg/ml/peptide). Internal plate controls were R10 alone (without cells) and anti-CD3 (Mabtech mAb CD3-2). At the end of the stimulation period, cell culture supernatants were collected and stored for later cytokine analysis by Luminex. ELISpot plates were developed with human biotinylated IFNγ detection Ab, directly conjugated to alkaline phosphatase (7-B6-1-ALP, Mabtech; 1µg/ml), diluted in PBS with 0.5% FCS, incubating 50µl/well for 2h at room temperature. This was followed by 50µl/well of sterile filtered BCIP/NBT-plus Phosphatase Substrate (Mabtech) for 5 minutes at room temperature. Plates were washed in ddH20 and left to dry completely before being read on AID-ELISpot plate reader. For experiments involving T cell stimulation with OLP peptide pools and Spike MEP2 pool ELISpot plates (AID classic ELISpot plate reader (Autoimmun Diagnostika GMBH, Germany) were coated with human anti-IFNγ Ab (1-D1K, Mabtech; 10µg/ml) in PBS overnight at 4°C. Plates were washed x6 with sterile PBS and were blocked with R10 for 2h at 37°C with 5% CO_2_. PBMC were thawed and rested in R10 for 3h at 37 °C with 5%CO_2_ before being counted. 400,000 PBMC were seeded in R10/well and were stimulated for 16-20h with SARS-CoV-2 OLP pools or Spike MEP2 pool (2µg/ml/peptide). Internal plate controls were R10 alone (without cells) and two DMSO wells (negative controls), concanavalin A (ConA, positive control; Sigma-Aldrich) and FEC (HLAI-restricted peptides from influenza, Epstein-Barr virus, and CMV; 1µg/ml). ELISpot plates were developed with human biotinylated IFN-γ detection antibody (7-B6-1, Mabtech; 1µg/ml) for 3h at room temperature, followed by incubation with goat anti-biotin alkaline phosphatase (Vector Laboratories; 1:1000) for 2h at room temperature, both diluted in PBS with 0.5% BSA by volume (Sigma-Aldrich), and finally with 50µl/well of sterile filtered BCIP/NBT Phosphatase Substrate (ThermoFisher) for 7 minutes at room temperature. Plates were washed in ddH20 and left to dry overnight before being read on an AID classic ELISpot plate reader (Autoimmun Diagnostika GMBH, Germany).

Analysis of ELISpot data was performed in Microsoft Excel. The average of two R10 alone wells or DMSO (Sigma-Aldrich) wells was subtracted from all peptide stimulated wells and any response that was lower in magnitude than 2 standard deviations of the sample specific control wells was not considered a peptide specific response. Results were expressed as difference in (delta) spot forming cells per 10^6^ PBMC between the negative control and protein/peptide stimulation conditions. We excluded the results if negative control wells had >100 SFU/10^6^ PBMC or positive control wells (ConA or anti-CD3) were negative. Results were plotted using Prism v. 7.0e and 8.0 for Mac OS (GraphPad).

### Cytokine measurement

Concentrations of IL-4, IL-5, IL-13, IL-17a and IL-23 were measured by multiplex Luminex® assay (Bio-Techne) on a Bio-Plex 200 instrument (Bio-Rad Laboratories, Ltd). Cytokine levels in cell culture supernatants in response to PBMC stimulation with Spike or NP protein were calculated in Microsoft Excel by subtracting values obtained for media only controls. Standard curves were plotted using Prism 8.0 for Mac OS (GraphPad).

### Cell Lines

HEK-293T and Huh7 (both ATCC) were cultured and maintained in high glucose Dulbecco’s Modified Eagle’s Medium and supplemented with GlutaMAX, 10% (*v/v*) heat-inactivated foetal bovine serum (FBS, 56°C for 30 minutes), 100IU/ml penicillin and 100µg/ml streptomycin. Cell lines were cultured at 37°C with 5% CO_2_.

### Production and titration of SARS-CoV-2 pseudotyped lentiviral reporter particles

Pseudotype stocks were prepared by linear polyethylenimine 25K (Polysciences) co-transfection of HEK-293T (ATCC) with SARS-CoV-2 spike pcDNA expression plasmid, HIV gag-pol p8.91 plasmid and firefly luciferase expressing plasmid pCSFLW at a 1:1:1.5 ratio^37,38^. 2.5⨯10^4^ cells/cm^2^ were plated 24h prior to transfection in 60cm^2^ cell culture dishes. 48 and 72h post transfection, pseudotype-containing culture medium was harvested and centrifuged at 500xg for 5 minutes to clear cell debris. Aliquots were stored at -80°C. TCID assays were performed by transduction of Huh7 cells to calculate the viral titre and infectious dose for neutralisation assays. p24 ELISA was also used to determine input concentration.

### p24 ELISA

Pseudotype stock concentrations were determined by ELISA for p24 protein concentration as previously described^39^. White ELISA plates were pre-coated with 5µg/ml sheep anti-HIV-1 p24 antibody (Aalto Bio Reagents) at 4°C overnight. Pseudoviral supernatants were treated with 1% Empigen BB (Merck) for 30 min at 56°C and then plated at 1:10 dilution in Tris-buffered saline (TBS) on pre-coated plates and incubated for 3h at room temperature. Alkaline phosphatase-conjugated mouse anti-HIV-1 p24 monoclonal antibody (Aalto Bio Reagents) in TBS, 20% (*v/v*) sheep serum, 0.05% (*v/v*) Tween 20 was then added and incubated for 1h at room temperature. After 4 washes with phosphate-buffered saline (PBS)-0.01% (*v/v*) Tween 20 and 2 washes with ELISA Light washing buffer (ThermoFisher), CSPD substrate with Sapphire II enhancer (ThermoFisher) was added and incubated for 30 minutes at room temperature before chemiluminescence detection using a CLARIOStar Plate Reader (BMG Labtech).

### Pseudotyped SARS-CoV-2 neutralisation assays

SARS-CoV-2 pseudotype neutralisation assays were conducted using pseudotyped lentiviral particles as previously described^37-40^. Serum was heat-inactivated at 56°C for 30 minutes to remove complement activity. Serum dilutions in DMEM were performed in duplicate in white, flat-bottom 96-well plates (ThermoFisher, #136101) with a starting dilution of 1 in 20 and 7 consecutive 2-fold dilutions to a final dilution of 1/2,560 in a total volume of 100µl. 1 ⨯10^5^ RLU of SARS-CoV-2 pseudotyped lentiviral particles were added to each well and incubated at 37°C for 1h. 8 control wells per plate received pseudotype and cells only (virus control) and another 8 wells received cells only (background control). 4⨯10^4^ Huh7 cells suspended in 100μl complete media were added per well and incubated for 72h at 37°C and 5% CO_2_. Firefly luciferase activity (luminescence) was measured using Steady-Glo® Luciferase Assay System (Promega) and a CLARIOStar Plate Reader (BMG Labtech). The curves of relative infection rates (in %) versus the serum dilutions (log10 values) against a negative control of pooled sera collected prior to 2016 (Sigma) and a positive neutraliser were plotted using Prism 8 (GraphPad). A non-linear regression method was used to determine the dilution fold that neutralised 50% (IC50).

### Statistics and reproducibility

Data was assumed to have a non-Gaussian distribution. Non-parametric tests were used throughout. For single paired and unpaired comparisons Wilcoxon matched-pairs signed rank test and a Mann-Whitney t-test were used. For multiple paired and unpaired comparisons Friedman multiple comparisons ANOVA with Dunn’s correction or Kruskal-Wallis one-way Anova with Dunn’s correction were used. For correlations, Spearman’s r test was used. A p value <0.05 was considered significant. Prism v. 7.0e and 8.0 for Mac was used for analysis.

## Acknowledgements

The authors wish to thank all the HCW participants for donating their samples and data for these analyses, and the research teams involved in consenting, recruitment and sampling of the HCW participants. Funding for COVIDsortium was donated by individuals, charitable Trusts, and corporations including Goldman Sachs, Citadel and Citadel Securities, The Guy Foundation, GW Pharmaceuticals, Kusuma Trust, and Jagclif Charitable Trust, and enabled by Barts Charity with support from UCLH Charity. Wider support is acknowledged on the COVIDsortium website. Institutional support from Barts Health NHS Trust and Royal Free NHS Foundation Trust facilitated study processes, in partnership with University College London and Queen Mary University London. We thank Antonio Bertoletti for supplying pre-pooled OLPs and spike MEP2. RJB/DMA are supported by MRC Newton (MR/S019553/1 and MR/R02622X/1), NIHR Imperial Biomedical Research Centre (BRC):ITMAT, Cystic Fibrosis Trust SRC, and Horizon 2020 Marie Curie Actions. MKM is supported by the UKRI/NIHR UK-CIC grant, a Wellcome Trust Investigator Award (214191/Z/18/Z) and a CRUK Immunology grant (26603). LS is supported by a Medical Research Foundation fellowship (044-0001). ÁM is supported by Rosetrees trust, The John Black Charitable Foundation, and Medical College of St Bartholomew’s Hospital Trust. JCM, CM and TAT are directly and indirectly supported by the University College London Hospitals (UCLH) and Barts NIHR Biomedical Research Centres and through the British Heart Foundation (BHF) Accelerator Award (AA/18/6/34223). TAT is funded by a BHF Intermediate Research Fellowship (FS/19/35/34374). MN is supported by the Wellcome Trust (207511/Z/17/Z) and by NIHR Biomedical Research Funding to UCL and UCLH.

The funders had no role in study design, data collection, data analysis, data interpretation, or writing of the report.

## Author contributions

R.J.B, D.M.A, M.K.M and Á.M. conceptualised the research project reported. R.J.B., D.M.A, and M.K.M supervised the T cell experiments. Á.M. supervised the nAb experiments. T.B. and A.Se supervised S1 IgG and NP IgG/IgM studies. C.J.R., D.B., L.S., N.S., M.D., and O.A. performed and analysed the T cell experiments. J..M.G. and C.P. performed and analysed the nAb experiments. C.G. and N.T. developed Pseudotyped SARS-CoV-2 neutralisation assays. S.T. performed and J.J. and A.Se analysed the S1 IgG and NP IgG/IgM assays. T.B., C.M., Á.M., T.T., J.M., and M.N. conceptualised and established the HCW cohort. M.J. analysed the HCW database. R.J.B., T.T., C.M., J.M., M.N., and M.J. designed the 16-18 week sub-study recruitment. M.J., W.L., M.J., J.R., A.C., G.J., J.L., M.F., A.S., C.M., T.T., and J.M. collected HCW samples and data, established the HCW cohort data base. J.M.G., C.P., C.J.R., D.B., S.B., L.S., N.S., M.D., O.A., J.R., and A.C. processed HCW samples. B.O’B. provided critical reagents. R.J.B., M.J., C.J.R, S.B., L.S., J.M.G., and C.P. analysed the data. R.J.B., M.K.M., Á.M., and D.M.A. wrote the manuscript. C.J.R., L.S., J.M.G., C.P., and M.J. helped prepare the manuscript and figures. All the authors reviewed and edited the manuscript and figures.

---

**Extended Data Figure 1.**
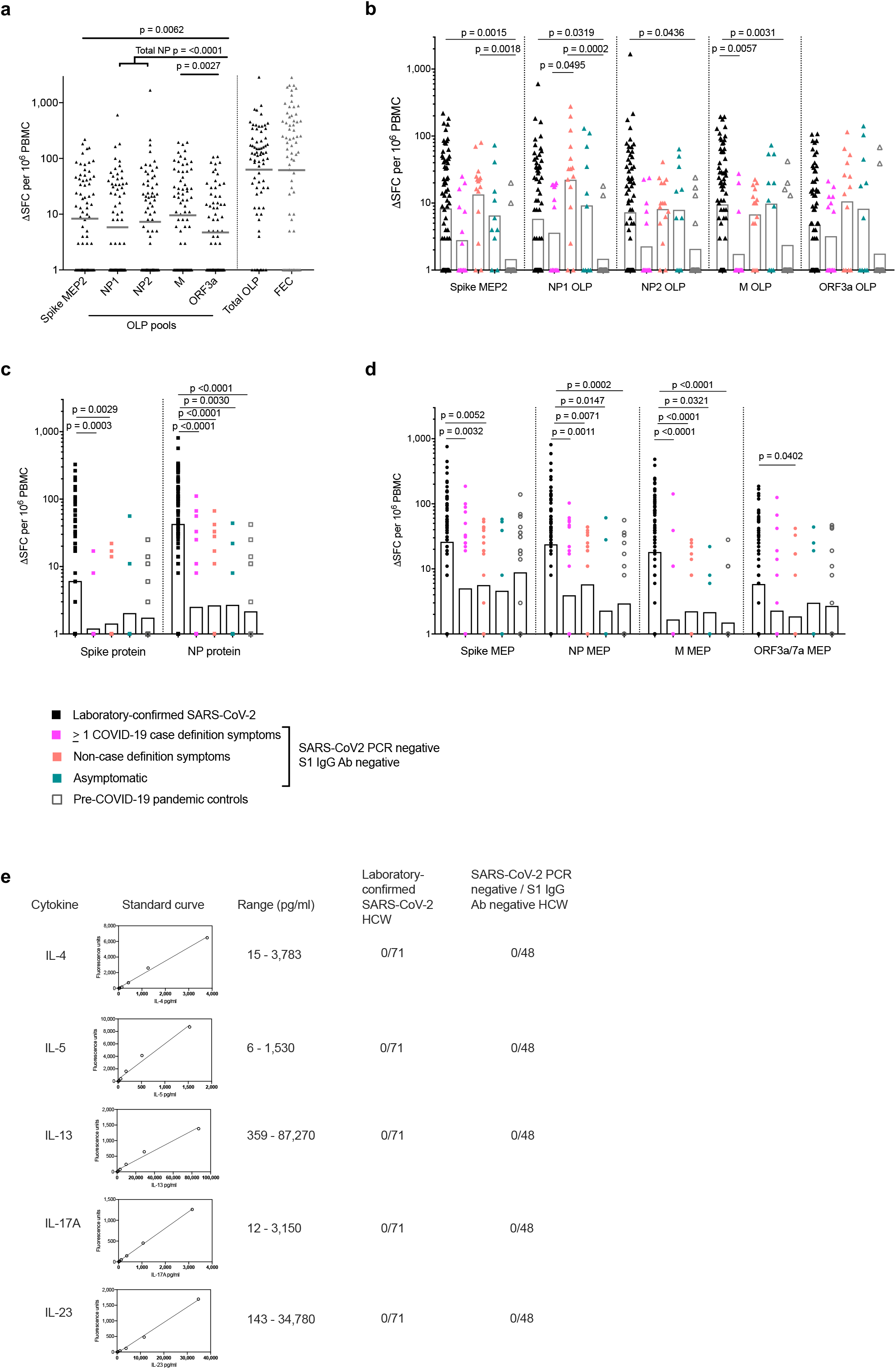
T cell responses to SARS-CoV-2 antigens in HCW with and without laboratory-confirmed SARS-CoV-2 infection 16-18 weeks after UK lockdown: **a)** Magnitude of the T cell response to Spike mapped epitope pool 2 (Spike MEP2), overlapping peptide pools (OLP), the summed total response to OLP pools + Spike MEP2, and response to positive control FEC peptide pool (covering CD8 epitopes from flu, EBV, CMV) in HCW with laboratory-confirmed SARS-CoV-2 infection (n = 71). **b)** Magnitude of the T cell response to Spike MEP2 and OLP pools in the following groups: HCW with laboratory-confirmed SARS-CoV-2 infection (n = 71); HCW with no laboratory-confirmed infection but with ≥1 COVID-19 case definition symptoms (n = 15), non-case definition symptoms (n = 15) or asymptomatic (n = 10); pre-COVID-19 pandemic control cohort 2 (n = 19). **c**,**d)** Magnitude of the T cell response to Spike and NP proteins **c)** and mapped epitope pools (MEP) **d)** in the following groups: HCW with laboratory-confirmed SARS-CoV-2 infection (n = 75); HCW with no laboratory-confirmed infection but with ≥1 COVID-19 case definition symptoms (n = 26), non-case definition symptoms (n = 24) or asymptomatic (n = 9); pre-COVID-19 pandemic control cohort 1 (n = 20). **e)** Number of HCW with detectable IL-4, IL-5, IL-13, IL-17A or IL-23 cytokine levels in T cell ELISpot supernatants by Luminex assay. Example cytokine standard curves and the concentration range of each cytokine assay are shown. **a-d)** Bars at geomean. **a)** Friedman multiple comparisons ANOVA with Dunn’s correction. **b-d)** Kruskal-Wallis multiple comparisons ANOVA with Dunn’s correction. Ab, antibody; HCW, health care workers; M, Membrane; ORF, open reading frame; NP, Nucleoprotein; S1, Spike subunit 1; SFC, spot forming cells per 10^6^ PBMC.

**Extended Data Figure 2.**
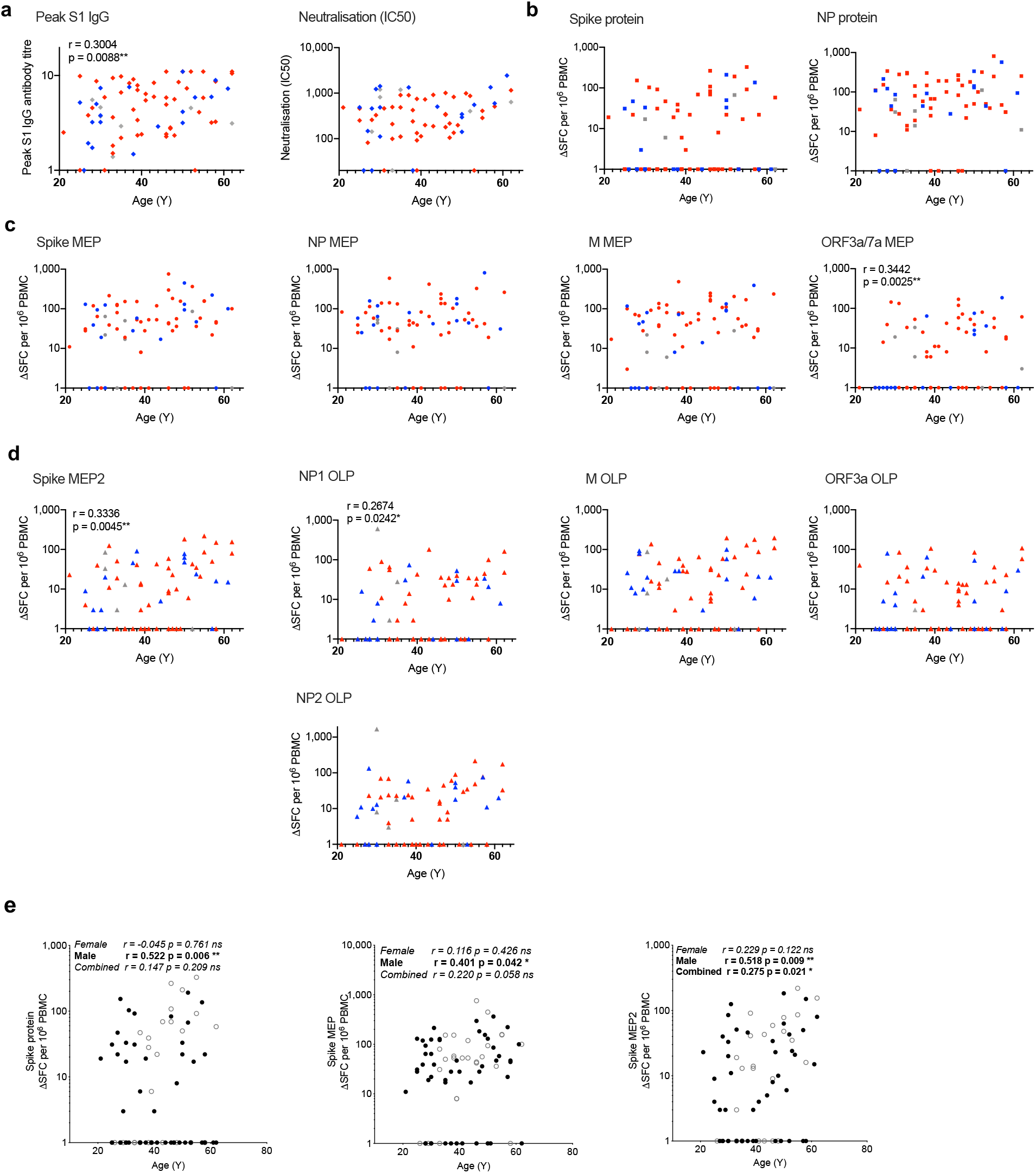
SARS-CoV-2 specific immune responses in HCW with laboratory-confirmed infection by age: **a-d)** Correlations between the peak S1 IgG antibody titre (left) and nAb titre (IC50; right) **a)**, T cell response to individual Spike and NP proteins **b)**, mapped epitope peptide pools (MEP) **c)** or Spike mapped epitope peptide pool 2 (MEP2) and overlapping peptide pools (OLP) **d)** vs. age, coloured by symptom group, in HCW with laboratory-confirmed SARS-CoV-2 infection (n = 71-75): Red, ≥1 case-definition symptom; Blue, ≥1 non-case definition symptom; Grey, asymptomatic throughout trial and within 3 months of trial initiation. **e)** Correlations of age (Y) vs. T cell response to Spike protein (left), Spike MEP (middle), Spike MEP2 (right) in HCW with laboratory-confirmed SARS-CoV-2 infection (Spike protein, Spike MEP, n = 75; Spike MEP2, n = 71) separated by gender (female, black circles; male, open circles). **a-e)** Spearman’s rank correlation. Ab, antibody; combined, correlation including both male and female HCW; HCW, health care workers; M, Membrane; ORF, open reading frame; ns, not significant; nAb, neutralising antibody; NP, Nucleoprotein; S1, Spike subunit 1; SFC, spot forming cells per 10^6^ PBMC; Y, years.

**Extended Data Figure 3.**
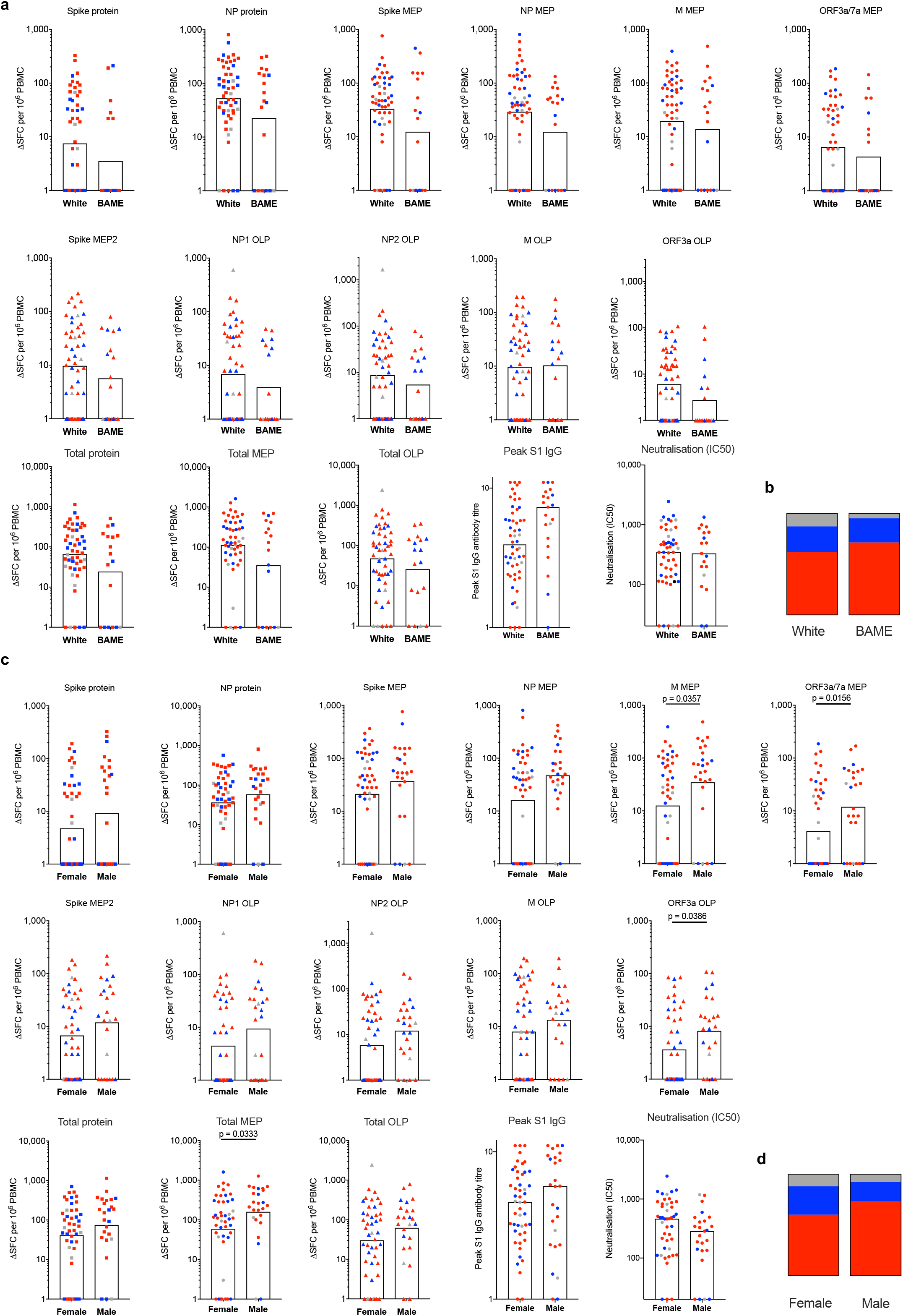
SARS-CoV-2 specific immune responses in HCW with laboratory-confirmed infection by gender and ethnicity: **a)** Magnitude of T cell responses to individual SARS-CoV-2 proteins, MEP and OLP pools, Peak S1 IgG Ab titre and nAb titre (IC50) in white and BAME HCW with laboratory-confirmed SARS-CoV-2 infection (n = 76). **b)** Proportion of white and BAME HCW with laboratory-confirmed SARS-CoV-2 infection who had ≥1 COVID-19 case definition symptoms (Red), non-case defined symptoms (Blue) or were asymptomatic (Grey) (n = 76). **c)** Magnitude of T cell responses to individual SARS-CoV-2 proteins, MEP and OLP pools, Peak S1 IgG Ab titre and nAb titre (IC50) in female and male HCW with laboratory-confirmed SARS-CoV-2 infection (n = 76). **d)** Proportion of female and male HCW with laboratory-confirmed SARS-CoV-2 infection who had ≥ 1 COVID-19 case-definition symptoms (Red), non-case-defined symptoms (Blue) or were asymptomatic (Grey) (n = 76). **a**,**c)** Mann-Whitney t-test. BAME, Black, Asian and Minority Ethnic; HCW, health care workers; M, Membrane; ORF, open reading frame; nAb, neutralising antibody; NP, Nucleoprotein; S1, Spike subunit 1; SFC, spot forming cells per 10^6^ PBMC.

**Extended Data Figure 4.**
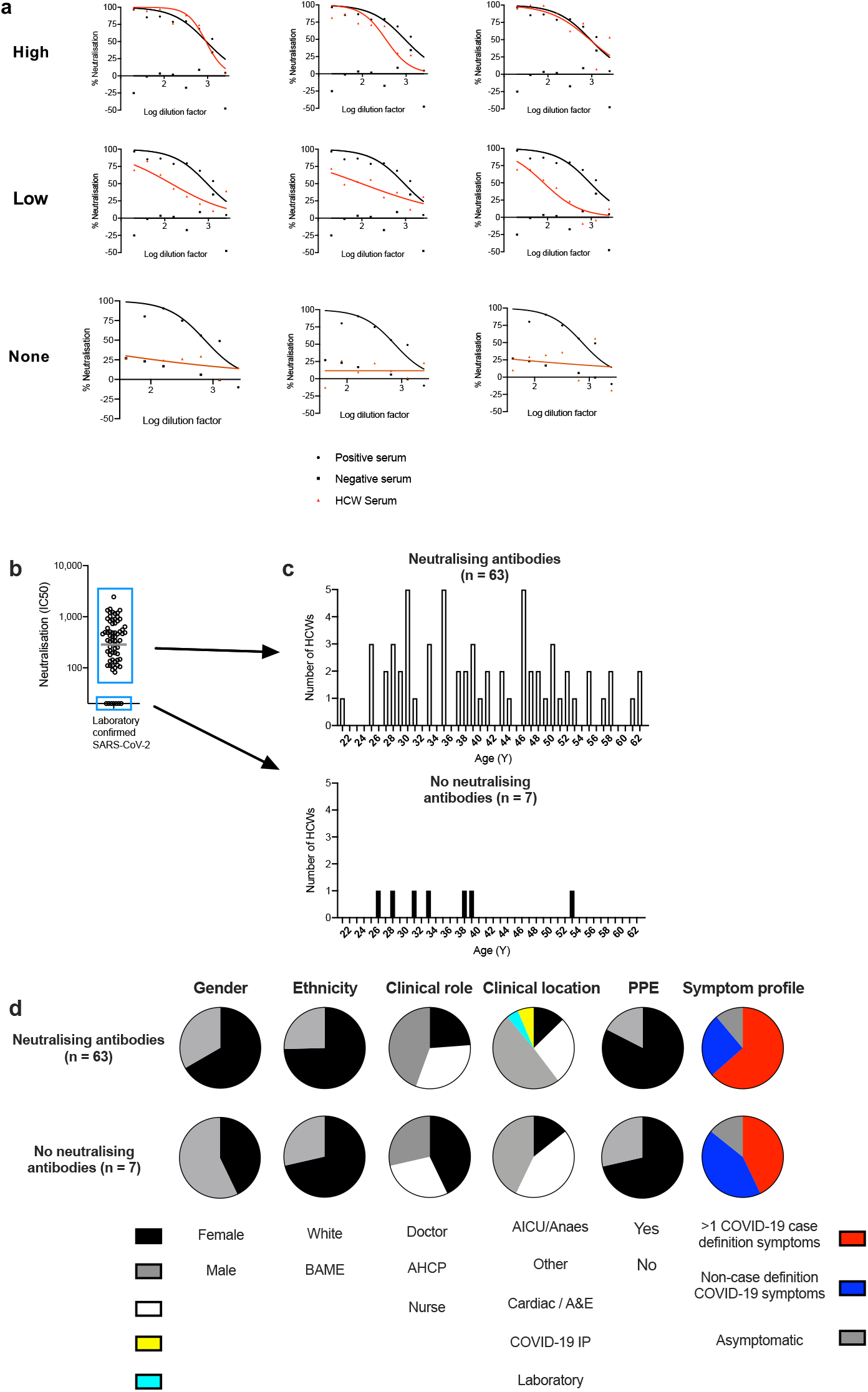
Demographic characteristics of HCW with laboratory-confirmed SARS-CoV-2 infection but no nAb at 16-18 weeks after UK lockdown. **a)** Neutralisation (%) curves of serum from selected HCW with laboratory-confirmed SARS-CoV-2 infection (red line with red triangles), a positive neutralising control serum (black line with black circles) and a negative non-neutralising control serum (black squares). Example neutralisation curves of participants with high (top row), low (middle row) or no neutralising ability (bottom row) are shown. **b)** The nAb titre (IC50) in HCW with laboratory-confirmed SARS-CoV-2 infection (n = 70). HCW with no neutralising antibody (n = 7) are within the lower blue box. **c)** The age in years of HCW for whom nAb were detected (top) or were not (bottom). **d)** Proportion of HCW with nAb (top row), or no nAb (bottom row) stratified by the demographic characteristics of gender, ethnicity, clinical role, clinical location, PPE or symptom profile. **e)** Bar at geomean. A&E, Accident and Emergency; AHCP, Allied health care professional; AICU, Adult intensive care unit; BAME, Black, Asian and Minority Ethnic; COVID-19 IP, COVID-19 in patient ward; HCW; health care workers; nAb, neutralising antibody; PPE, Personal protective equipment; Y, years.

**Extended Data Figure 5.**
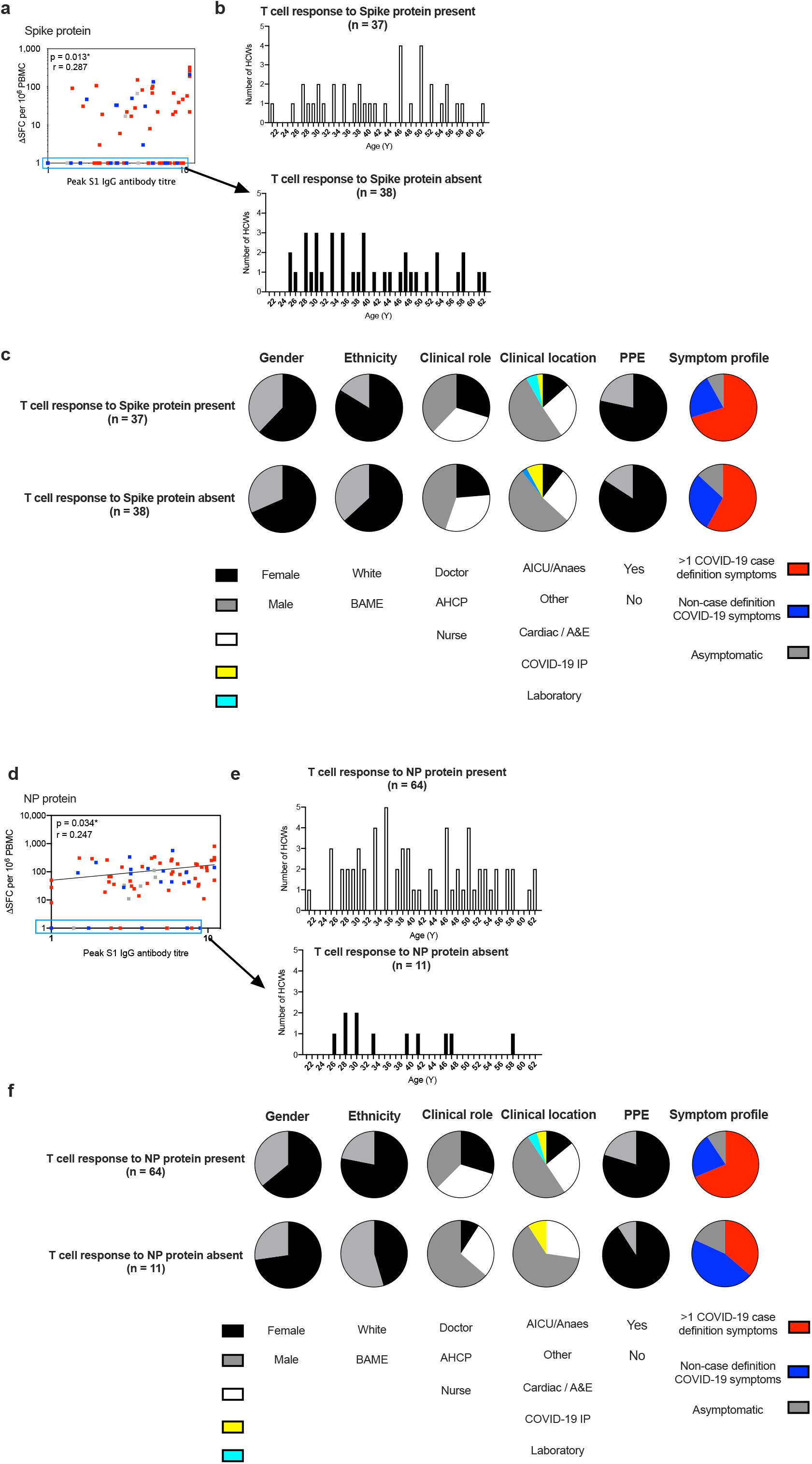
Demographic characteristics of health care workers with laboratory-confirmed SARS-CoV-2 infection but no T cell response to Spike or NP protein at 16-18 weeks after UK lockdown: **a)** Correlation between peak S1 IgG Ab titre and T cell response to Spike protein in HCW with laboratory-confirmed SARS-CoV-2 infection, coloured by symptom group: HCW who had ≥ 1 COVID-19 case definition symptoms (Red), non-case definition symptoms (Blue) or were asymptomatic (Grey). HCW with no T cell response to Spike protein are within the blue box. **b)** The age in years of HCW for whom a T cell response to Spike protein was detected (top) or was not (bottom). **c)** Proportion of HCW with a T cell response to Spike protein (top row), or no T cell response to Spike protein (bottom row) stratified by the demographic characteristics of gender, ethnicity, clinical role, clinical location, PPE or symptom profile. **d)** Correlation between peak S1 IgG Ab titre and T cell response to NP protein in HCW with laboratory-confirmed SARS-CoV-2 infection, coloured by symptom group as above. HCW with no T cell response to NP protein are within the blue box. **e)** The age in years of HCW for whom a T cell response to NP protein was detected (top) or was not (bottom). **f)** Proportion of HCW with a T cell response to NP protein (top row), or no T cell response to NP protein (bottom row) stratified by the demographic characteristics of gender, ethnicity, clinical role, clinical location, PPE or symptom profile. **a, d)** Spearman’s rank correlation. A&E, Accident and Emergency; AHCP, Allied health care professional; AICU, Adult intensive care unit; BAME, Black, Asian and Minority Ethnic; COVID-19 IP, COVID-19 in patient ward; PPE, Personal protective equipment; S1, Spike subunit 1; SFC, spot forming cells per 10^6^ PBMC; Y, years.

**Extended Data Figure 6.**
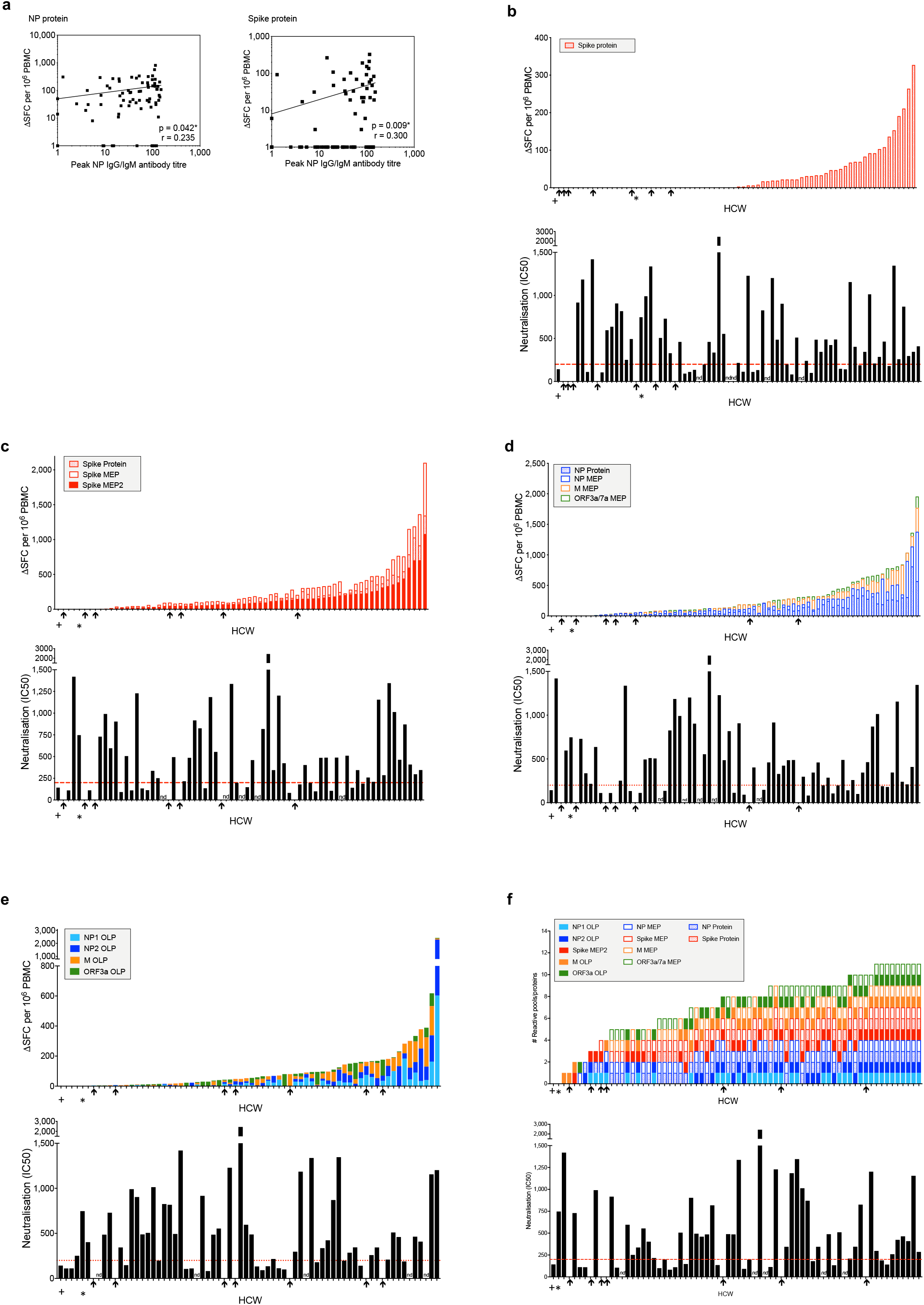
Correlations between antibody and T cell responses in HCW with laboratory-confirmed SARS-CoV-2 infection. **a)** Correlations between the peak NP IgG/IgM antibody titre and T cell responses to NP protein (left) and Spike protein (right) in HCW with laboratory-confirmed SARS-CoV-2 infection (n=75) **b-e)** Top panels; Magnitude of the T cell response to Spike protein (n = 75) **b)** Cumulative magnitude of T cell responses to Spike protein and Spike mapped epitope peptide (MEP and MEP2) pools (n = 70) **c)** NP protein and NP, M and ORF3a/7a MEP pools (n = 75) **d)** or NP1, NP2, M and ORF3a overlapping peptide (OLP) pools (n = 70) **e)** ordered by increasing cumulative magnitude of T cell responses in HCW with laboratory-confirmed SARS-CoV-2 infection. Bottom panels; nAb titres (IC50) in HCW with laboratory-confirmed SARS-CoV-2 infection, ordered by corresponding top panel. **f)** The number of reactive SARS-CoV-2 proteins or peptide pools (top panel) and nAb titre (IC50; bottom panel) in HCW with laboratory-confirmed SARS-CoV-2 infection (n = 70). Top panel ordered by cumulative magnitude; bottom panel ordered by top panel. HCW with no nAb (IC50 titre less than 50) are indicated by black arrows. + and * denote two individuals with no T cell response to any protein or peptide pool. **a)** Spearman’s rank correlation, least squares log-log lines shown. HCW, health care workers; M, Membrane; nd, not done; nAb; neutralising antibody; NP, Nucleoprotein; ORF, open reading frame; SFC, spot forming cells per 10^6^ PBMC.

**Extended Data Figure 7.**
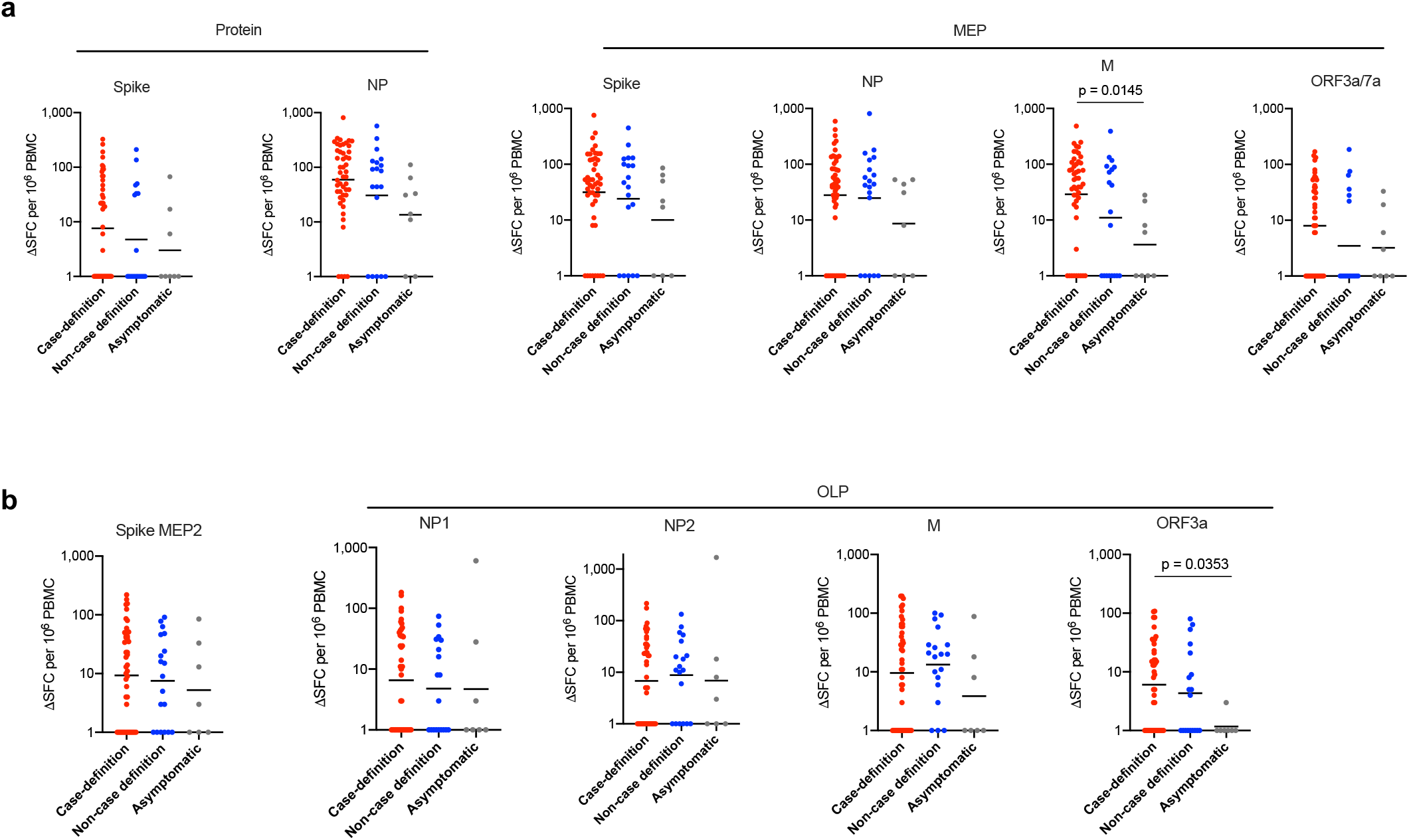
T cell responses to SARS-CoV-2 in HCW with laboratory-confirmed infection stratified by symptoms: **a)** T cell responses to Spike and NP proteins and to mapped epitope peptide pools (MEP) in HCW with laboratory-confirmed SARS-CoV-2 infection (n = 75) stratified by symptom group: ≥1 case-definition symptom (Red; n = 48); ≥1 non-case definition symptom (Blue; n = 19) or asymptomatic (Grey; n = 8) throughout trial and within 3-months of trial initiation. **b)** T cell responses to Spike MEP2 and overlapping peptide pools (OLP) in HCW with laboratory-confirmed SARS-CoV-2 infection (n = 71) stratified by symptom group: ≥1 case-definition symptom (Red; n = 45); ≥1 non-case definition symptom (Blue; n = 19) or asymptomatic (Grey; n = 7) throughout trial and within 3 months of trial initiation. **a**,**b)** Bars at geomean, Kruskal-Wallis multiple comparison ANOVA with Dunn’s correction. HCW, health care workers; M, Membrane; ORF, open reading frame; NP, Nucleoprotein; SFC, spot forming cells per 10^6^ PBMC).

**Extended Data Figure 8.**
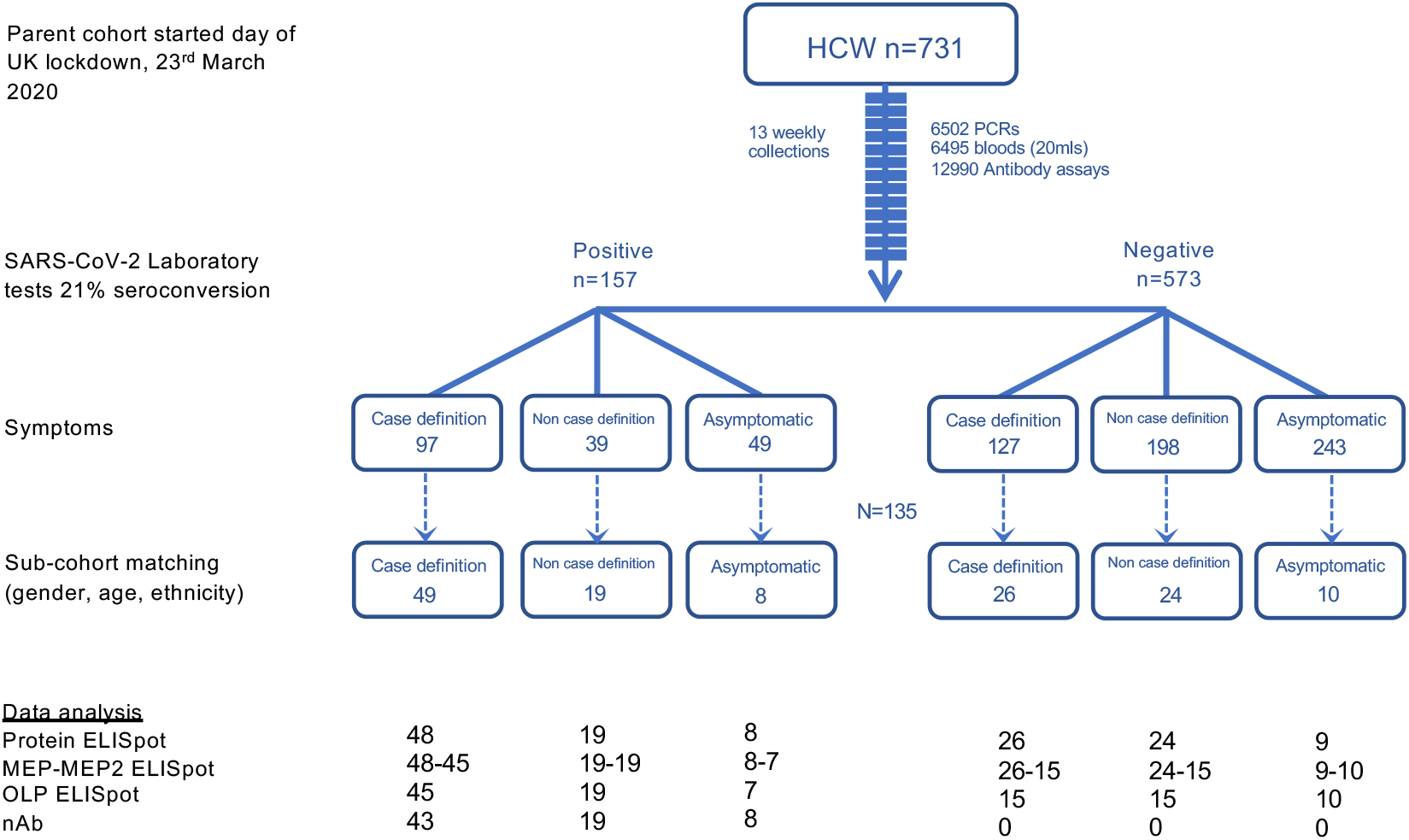
Consort flow diagram for the COVIDsortium London healthcare worker cohort and sub-cohort: CONSORT flow diagram showing participant recruitment into COVIDsortium London healthcare worker study. Participants were stratified by SARS-CoV-2 PCR and antibody laboratory tests and by symptoms experienced during follow-up and during the 3 months prior to study initiation. SARS-CoV-2 laboratory test positive and negative participant sub-cohort groups were matched for gender, age and ethnicity.

**Extended Data Table 1.**
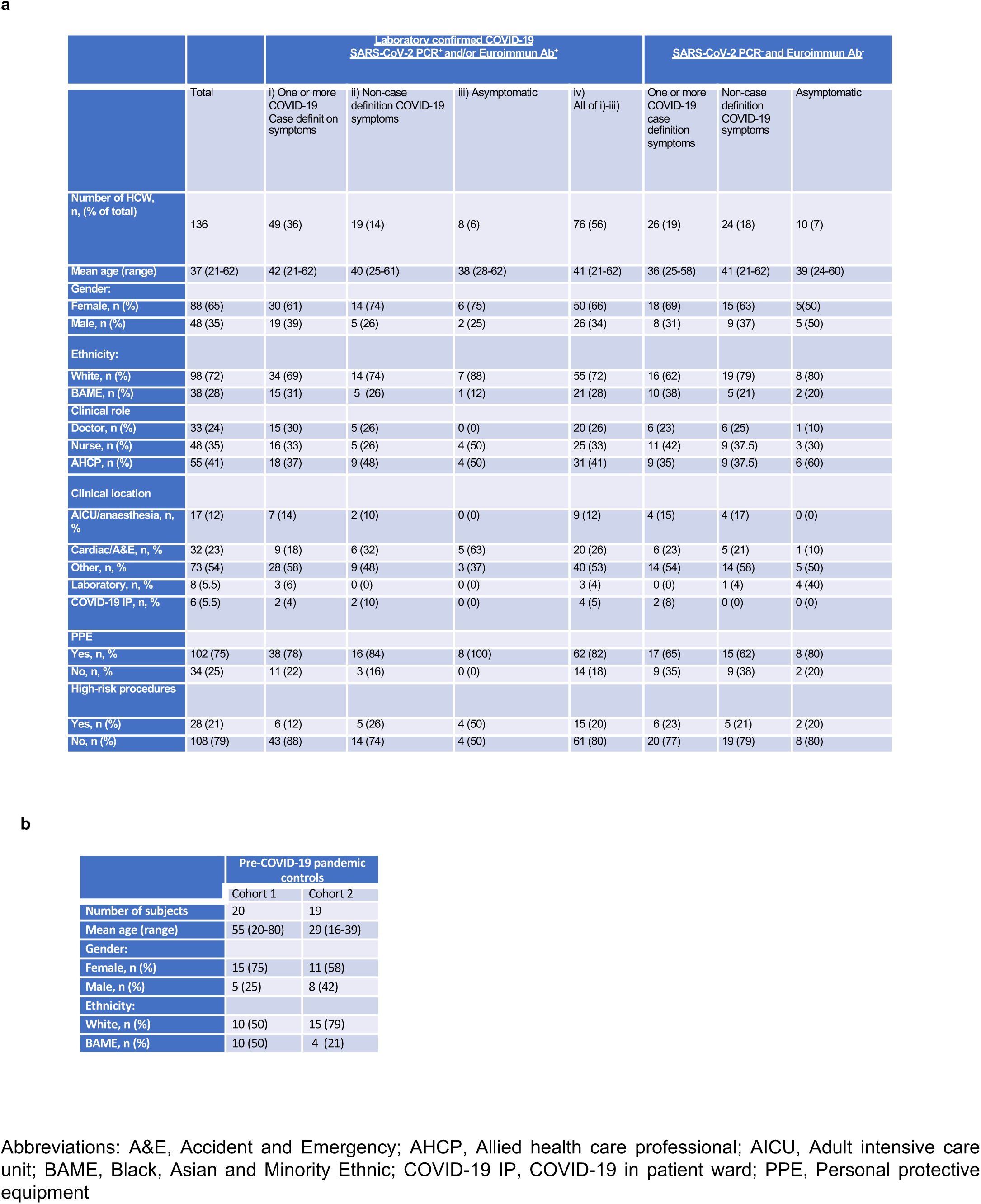
**a**, HCW characteristics and COVID-19 status. **b**, characteristics of pre-pandemic COVID-19 controls.

**Extended Data Table 2.**
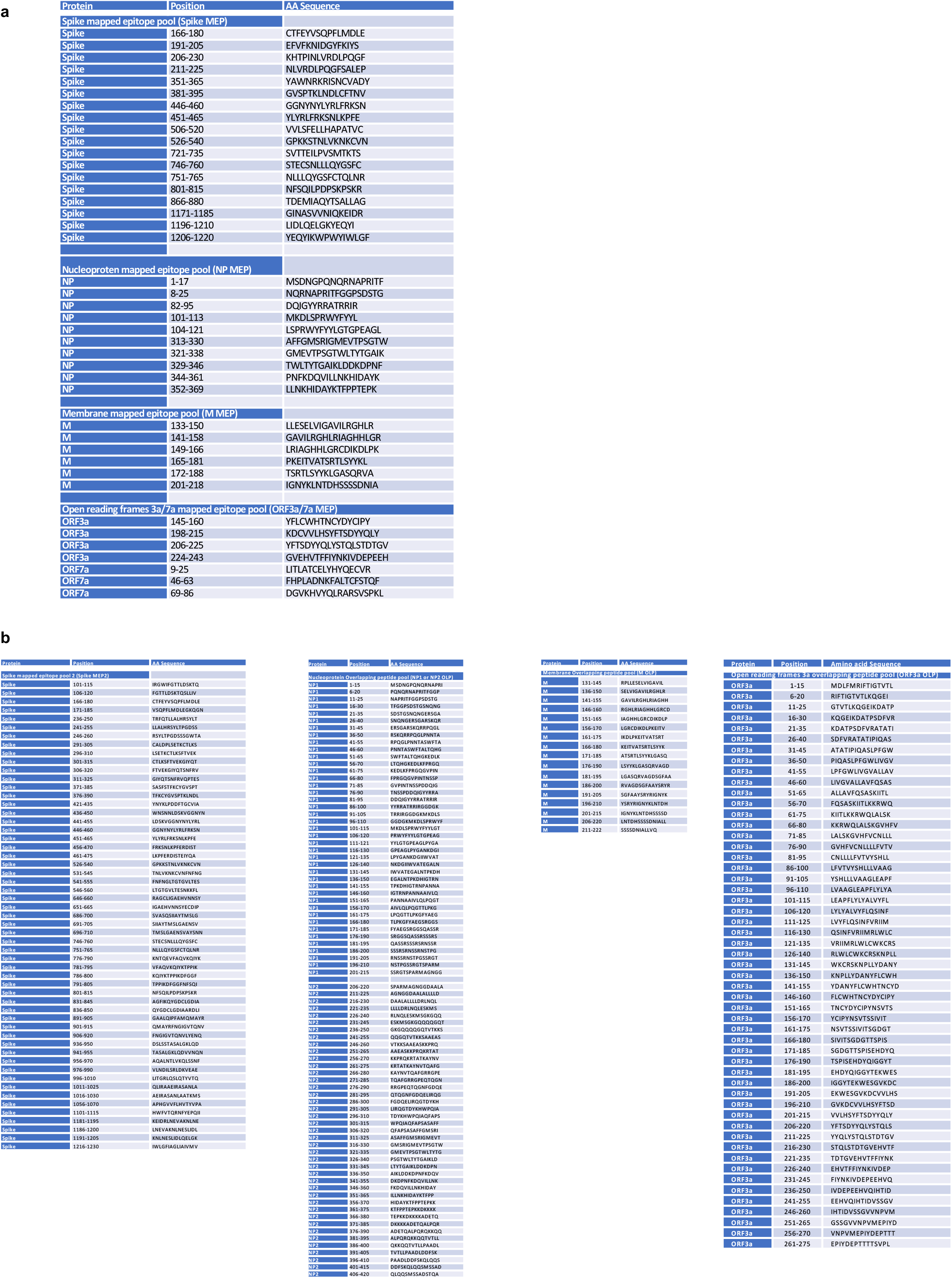
**a**, Mapped epitope peptide (MEP) pools. **b**, Spike MEP2 pool and overlapping peptides (OLP) pools.

